# Network Analysis of Pairwise Relative Tuberculosis Transmission Probabilities in Lima, Peru

**DOI:** 10.1101/2025.11.18.25340467

**Authors:** Anne N. Shapiro, Meredith B. Brooks, Chuan-Chin Huang, Megan B. Murray, Laura F. White, Helen E. Jenkins

## Abstract

**Background:** Identifying transmission events is important in understanding infectious disease dynamics. Such events are typically unobservable, particularly in diseases with long serial intervals such as tuberculosis (TB). We apply network techniques to identify transmission clusters and features shared within clusters.

**Methods:** We estimate directed pairwise transmission probabilities via an existing iterative algorithm that employs a modified Naïve Bayes classifier to incorporate demographic, clinical, and genetic data and use these probabilities to create a network. We explore noise reduction techniques to trim low probability edges. We apply clustering algorithms to group together individuals with TB based on edges informed by transmission probabilities. We apply our framework to simulated data and assess how the clustering algorithms captured the simulated clusters. We then apply this approach to data from a cohort study in Lima, Peru and examine the homogeneity of the clusters using a binary entropy measure.

**Results:** We find cluster performance to be consistent across all edge trimming scenarios and clustering methods. We find high levels of entropy for age, sex, socioeconomic status, and individuals who work outside the house and use public transit, indicating these variables are heterogenous across clusters.

**Conclusions:** We demonstrate approaches to analyze estimated directed pairwise transmission probabilities with network techniques. The approach is consistent across network construction and clustering methods. This method can be applied to any disease outbreak to understand its dynamics.

## 1. INTRODUCTION

Understanding transmission trends is critical to informing targeted interventions to interrupt infectious disease spread. However, transmission is often unobservable and difficult to trace, particularly for diseases with long serial intervals such as tuberculosis (TB).^1^ Despite TB being a leading cause of death globally^2^, its epidemiological parameters remain poorly informed, largely due to these tracing challenges.^3, 4^

Common methods to quantify transmission trends use genetic data, often single nucleotide polymorphisms (SNP) differences obtained from pathogen whole genome sequencing (WGS), to form transmission clusters. Isolates are included in a cluster if they are within a predetermined SNP distance from at least one other isolate in the cluster. ^5, 6, 7, 8^ By construction, two individuals in a cluster can be separated by more than the specified cutoff. This method has notable concerns: SNP distances alone cannot definitively determine transmission^9^ and results can vary depending on the selected SNP distance cutoff.^5^

In our previous work, henceforth called “mlTransEpi,” we estimated pairwise transmission probabilities using demographic and clinical data combined with SNP differences.^10^

mlTransEpi is a data driven method that does not assume that genetic differences alone can inform transmission events. Instead, it uses an iterative machine learning algorithm to estimate pairwise transmission probabilities. We seek to use these transmission probabilities to identify epidemiologically informed transmission clusters via a network graph, with each node being an individual with TB and each edge weighted by the directed transmission probability.

We hypothesize that the clusters identified from the transmission network improve on clusters created solely based on SNP distances. As stated earlier, SNP distances may not accurately represent transmission. Additionally, SNP clustering methods will add an individual to a cluster if they are within the cutoff of at least one other individual in the cluster; thus, not all individuals in the cluster are necessarily similar and/or share transmission events. In contrast, network-based clustering methods consider the relationship between all individuals within a cluster.

Here, we first examine the accuracy of these clustering methods via simulation. We then apply them to pairwise transmission probabilities estimated with published cohort data from Lima, Peru.^11^ We compare clusters created using network methods with mlTransEpi probabilities to those generated with traditional SNP cutoff methods. Finally, we analyze clusters to identify if the distributions of demographic and clinical features of individuals vary between clusters.

## 2. METHODS

### 2.1 Graph construction

We estimate directed pairwise transmission probabilities using mlTransEpi, described further elsewhere^10, 12, 13^ and shown in Figure 1.

**Figure 1.**
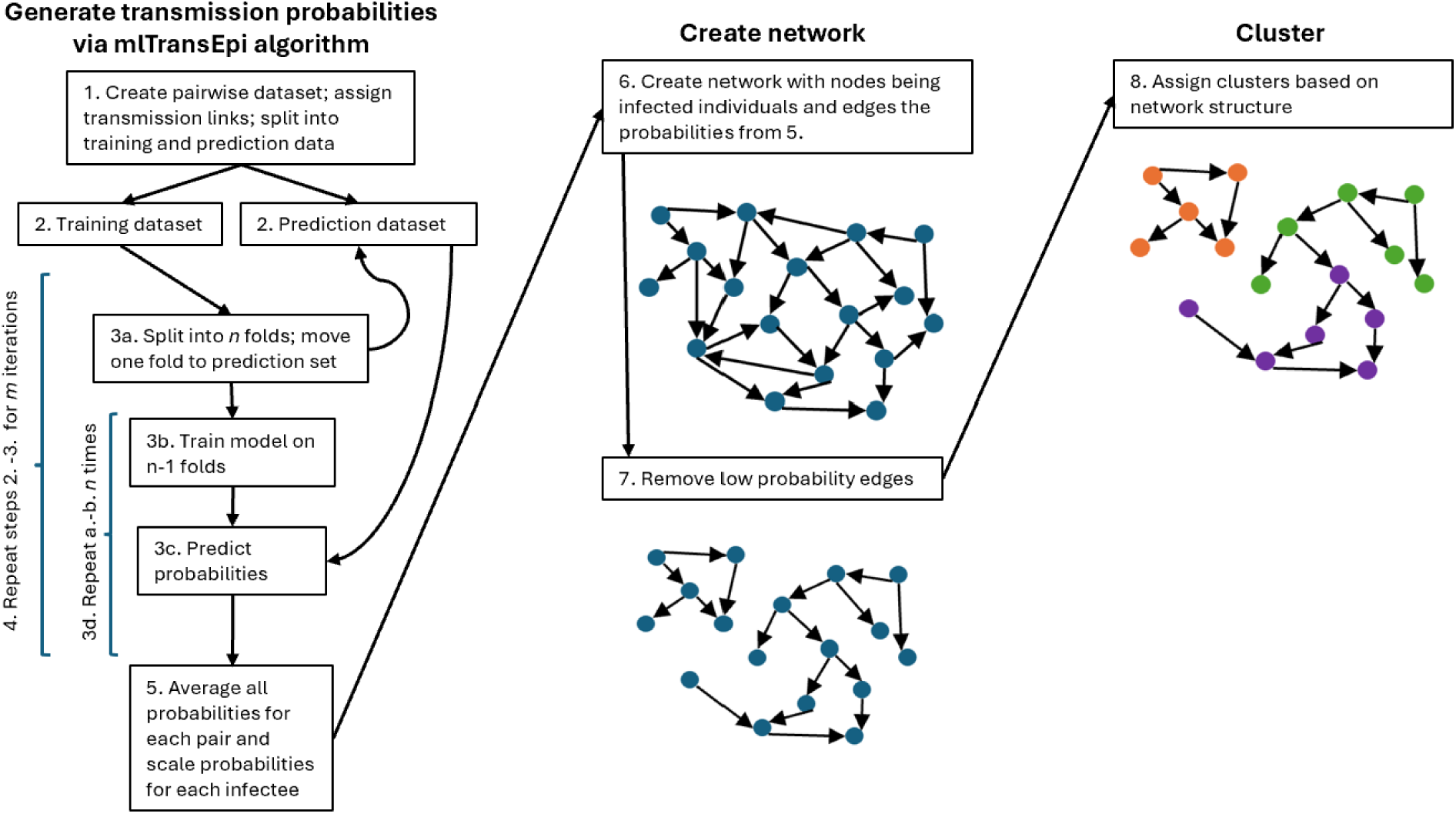
Diagram of the iterative process to generate transmission probabilities, create the network, and assign clusters.

We create a network graph where nodes are individuals with TB and each estimated transmission probability is a weighted, directed edge (where the weight is the estimated transmission probability). Our iterative algorithm estimates all pairwise probabilities; all pairwise edges between individuals will be nonzero, but many will be very small reflecting the extreme improbability that they are transmission links. To reduce noise, we consider three methods to remove these low probability edges: a probability cutoff, hierarchical clustering^14^, and kernel density estimation (see Supplementary materials S1 and Figure 1, panel 2).^15^

We examine two clustering methods: the Infomap algorithm^16^ and the Leiden algorithm.^17^ The calculation of modularity in the Leiden algorithm requires specification of a resolution parameter; higher resolution parameters result in more clusters and lower resolution lead to fewer clusters. There is no optimal value for the resolution parameter in the Leiden algorithm, rather it is selected based on the desired granularity of clusters. For this reason, we consider multiple values (see Figure 1, panel 3).^17^

### 2.2 Simulation study

We simulate 100 TB-like outbreaks following a procedure described in Supplementary Materials Section S2.

We apply the iterative algorithm to each of the simulated outbreaks with 10 iterations and 10 cross-validation folds. We consider pairs with fewer than 4 SNP differences to be probable links and pairs with greater than 12 SNP differences to be probable non-links. All pairs with 4-12 SNP differences are considered indeterminate and used only in the prediction dataset. We vary SNP difference boundaries in sensitivity analyses. We include all pairwise combinations in which the infector was observed before the infectee or up to one year after.

We assess edge trimming performance via multiple metrics. We calculate the number of true edges remaining as the number of edges corresponding to true transmission events not removed by our edge trimming method and sensitivity (Se), specificity (Sp), and positive predictive value (PPV) with the truth being an edge that corresponds to a true transmission instance.

We consider two types of true clusters to assess clustering performance. First, we allow each transmission chain to be a true cluster (henceforth “full outbreak”). Each simulation generates a random number of transmission chains. Second, we remove the first 12 years of each simulation and consider each separate subgraph (groups of nodes connected by transmission events) to be a true cluster (henceforth “trimmed outbreak”). In this scenario, we exclude nodes that become singletons (i.e. are not connected to any other node). This scenario is more similar to the data available from TB outbreaks, which are collected over a shorter period with the beginning of the epidemic unobserved. Figure 2 illustrates these two true cluster constructions.

**Figure 2.**
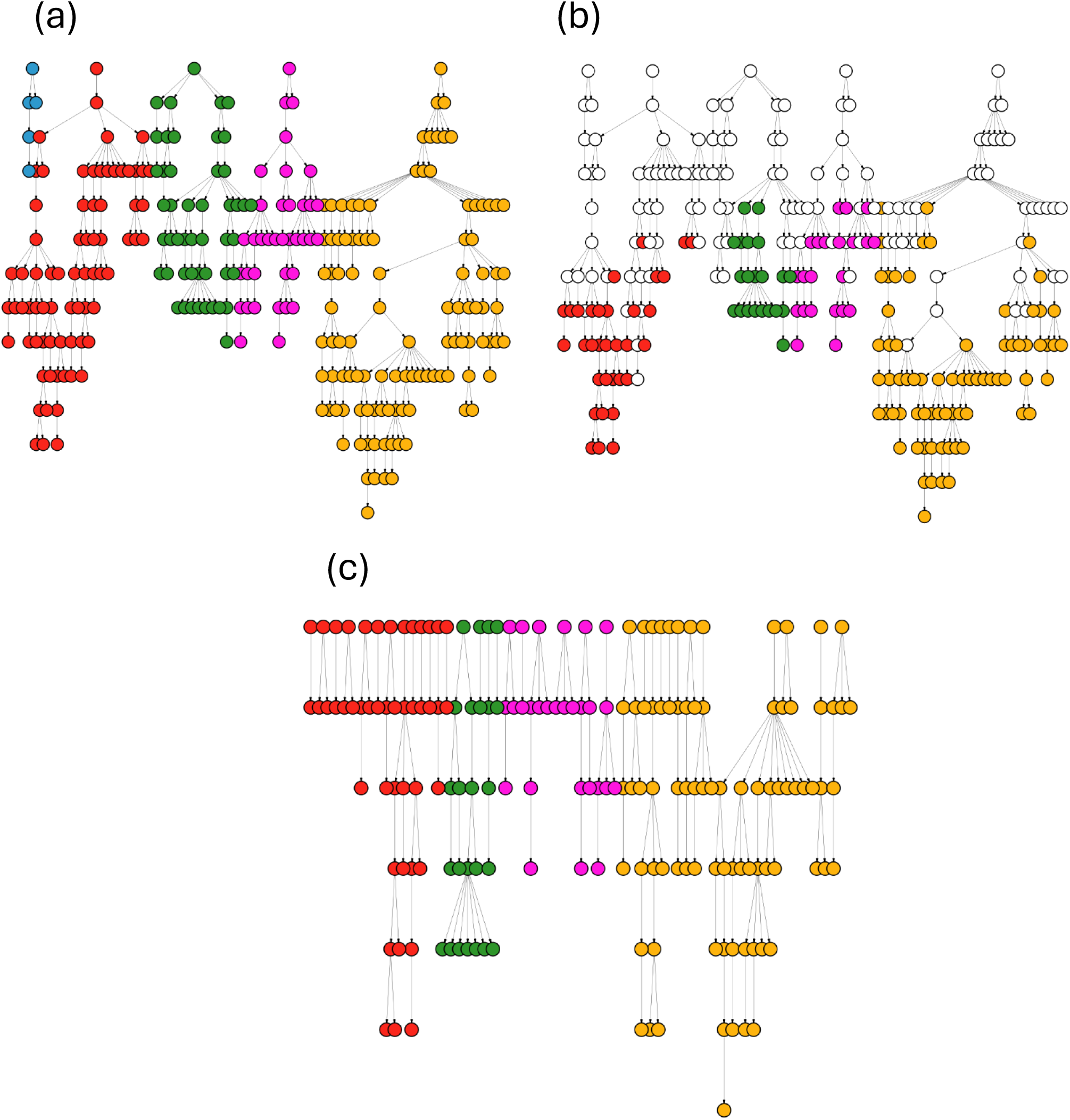
Single simulation example of true cluster assignments. In image (a), each of the transmission chains (differentiated by color) is considered a true cluster. Images (b) and (c) depict the process of creating true clusters after removing the first 12 years of each simulation. In image (b), all people diagnosed outside of the 8-year period of interest are colored white. Note that edge lengths and node positions do not scale to time. Image (c) depicts true clusters after removing people diagnosed after the first 12 years of our outbreak (with colors corresponding to their original outbreak chain); we consider each separate subgraph to be an outbreak.

We cluster using the Infomap algorithm and the Leiden algorithm with resolution parameters of 2, 3, and 4; note resolution is user-determined for the analytic goals of the network analysis.^18^ We assess cluster performance using the mutual information score (MI),^19^ pairwise F-score (F), and Fowlkes-Mallows index (FM).^20^ MI has a lower bound of 0 and no upper bound; F and FM are bounded between 0 and 1. A higher value of all three metrics indicates better clustering performance. See Supplementary Materials Section 2 for further details.

We perform a sensitivity analysis to assess clusters’ robustness to various sampling coverage scenarios. We examine four different scenarios: randomly sampling 50% and 80% of individuals (nodes), sampling only 80% of individuals in one class of the binary variable *X*_1_ (to simulate differential sampling by sex^21^), and individuals in one class of the categorical variable *X*_4_ (to simulate lack of access to healthcare in a specific neighborhood^22^). For each of these scenarios, we simulate an outbreak and remove individuals per the sampling scheme prior to calculating transmission probabilities. We also vary the time allowed between diagnosis of infectee and infector to assess the method’s robustness to diagnosis date misspecification.

### 2.3 Data application

We estimate pairwise transmission probabilities on data from a cohort study from Lima, Peru previously described by Trevisi et al.^11^ We consider all pairs in which a potential infector was diagnosed up to one year after a potential infectee and use the same SNP distance thresholds as in simulations. We use the pairwise variables as described in Trevisi et al.^11^ in our Naïve Bayes model. Individual-level and pairwise variables and their frequencies are described in Supplemental Materials Table S2a and S2b. We apply the iterative method with 60 iterations and 10 cross-validation folds.

We use hierarchical clustering with a cutoff of 0.1 to trim edges and the Infomap algorithm to create clusters for results in the main text; results using additional cutoffs and clustering methods are shown in the Supplementary Materials Section S3. We calculate the binary entropy of each cluster and calculate a weighted average by cluster size to assess the homogeneity of clusters. Binary entropy is a measure of the randomness of a binary variable. A value of 0 indicates perfect homogeneity and increasing values indicate increasing heterogeneity with the maximal value of 1 corresponding to equal probability of each feature of the binary variable within a cluster. We also calculate the proportion of perfectly homogenous clusters. We exclude clusters with fewer than 4 individuals from both homogeneity measures.

We also compare our methods to traditional SNP based clustering. For these clusters, we consider individuals to be in the same cluster if they were within the SNP distance cutoff of at least one other individual in the cluster. We also generate traditional clusters using SNP cutoffs of <12 and <20; commonly used cutoffs for SNP distance clustering.^5, 23^ We compare these to clusters generated using the mlTransEpi algorithm with an upper bound of 12 and 20 SNPs.

## 3. RESULTS

### 3.1 Simulation study

Edge trimming metrics are consistent for both hierarchical clustering cutoffs and kernel density estimation binwidths (Figure 3). Metrics using probability cutoffs have similar means to hierarchical clustering and kernel density estimation but larger variations (Figure 3). Clustering performance is also consistent across edge trimming methods for trimmed outbreak simulations (Figure 4). Clusters using the Leiden algorithm with a resolution parameter of 2 have slightly higher F and FM scores, indicating that this clustering algorithm best captures the true clusters for our simulated outbreaks.

**Figure 3.**
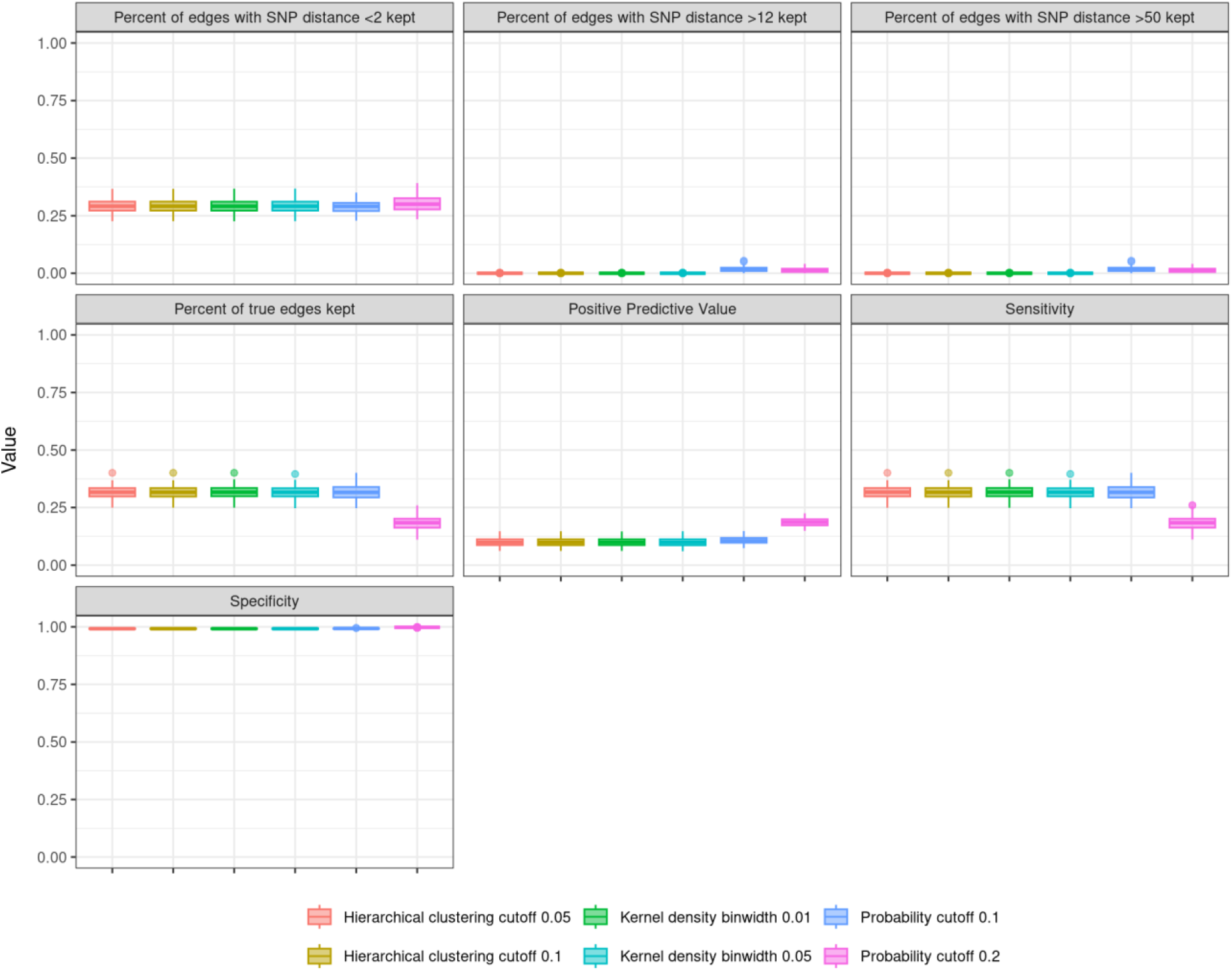
Edge trimming metrics from 100 simulated outbreaks, using the last 8 years of each outbreak. We consider three edge trimming methods: hierarchical clustering (HC), kernel density (KD) estimation, and probability cutoffs. Results are consistent across HC and KD edge trimming scenarios; probability cutoffs show more noise.

**Figure 4.**
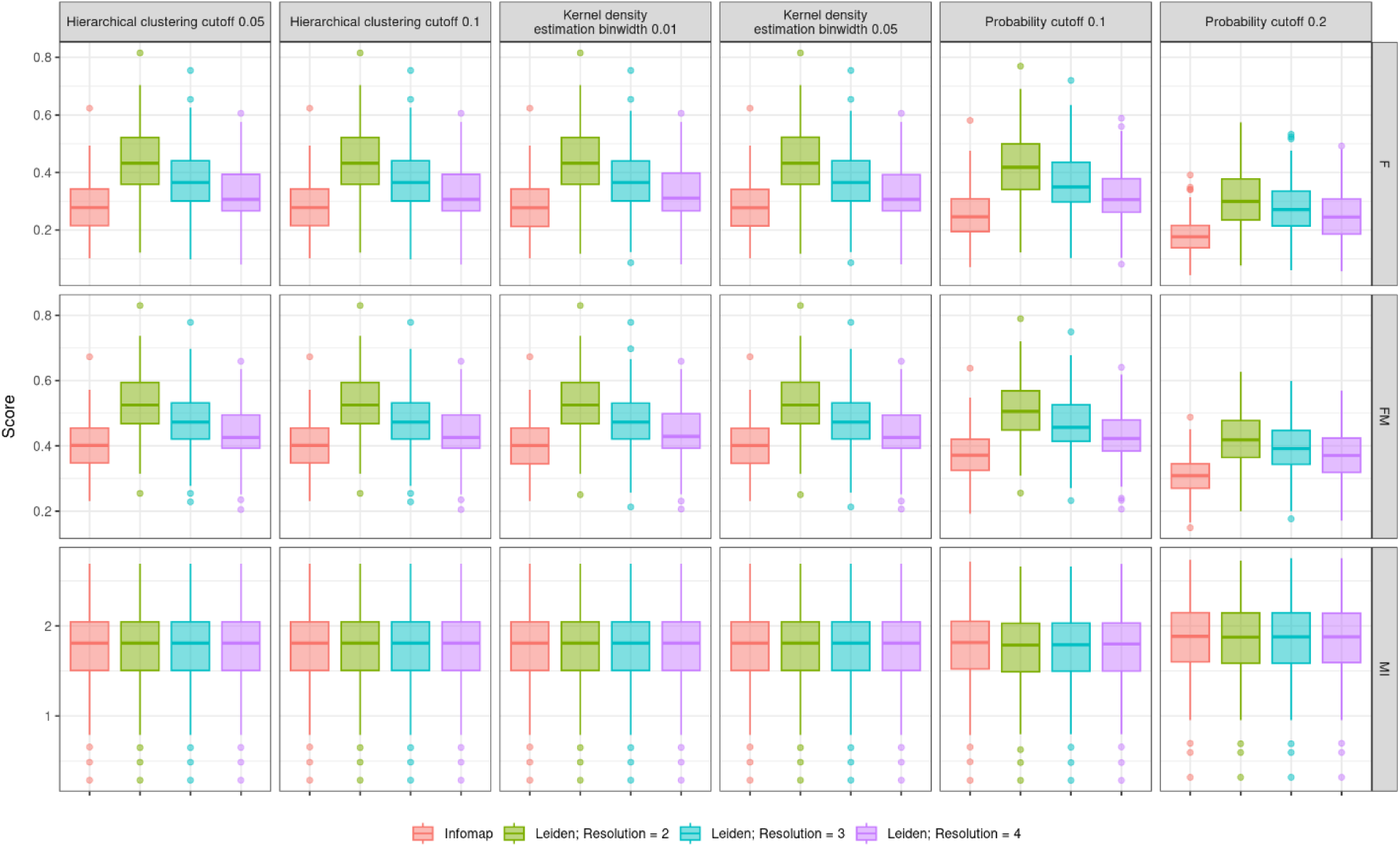
Pairwise F (F), Fowlkes-Mallows (FM), and mutual information (MI) scores assessing clustering performance for 100 simulated outbreaks, using the last 8 years of each outbreak. Results are consistent across all edge trimming scenarios.

These trends hold when using the full outbreak (Figures S1a and S1b). A SNP distance lower bound of 4 results in a higher percentage of true edges kept as well as F and FM scores, but trends are consistent across edge trimming and clustering methods for each set of bounds (Figures S2a and S2b). Similarly, positive predictive value increases as the generation interval distribution used to simulate the outbreak increases (Figures S3a and S3b) but trends also hold across edge trimming and clustering methods for each value.

Results are consistent when varying the amount of time allowed between diagnosis of the potential infector and infectee (Supplemental Materials Figure S4; recall that main results allow for the potential infector to be observed up to one year after the potential infectee) and across all sampling coverage scenarios (Supplemental Materials Figure S5).

### 3.2 Data application

Applying the Infomap clustering algorithm to a network trimmed with a hierarchical clustering cutoff of 0.1 results in 275 clusters. Of those, 163 clusters (59.2%) are of size 2 and 90% have 5 or fewer individuals. The largest cluster consists of 45 individuals. Network and cluster characteristics are similar for different hierarchical clustering cutoffs (used to trim low-probability edges; a higher cutoff will result in more edges being removed) and cluster methods (Table S3).

Variables for public transit use, working outside the home, age, sex, and socioeconomic status (SES) all have high weighted average entropy and low proportions of purely homogenous clusters (Figure 5). Recall that our entropy average is weighted by cluster size. HIV and drinking statuses have the lowest average entropy and highest proportion of homogeneity. Incarceration status also has a low average entropy. Supplemental materials Figure S6 shows individual binary entropy by variable for each cluster. Supplementary materials Figures S7 and S8 show the average entropy and proportion of homogeneity across multiple hierarchical clustering cutoffs and clustering methods.

**Figure 5.**
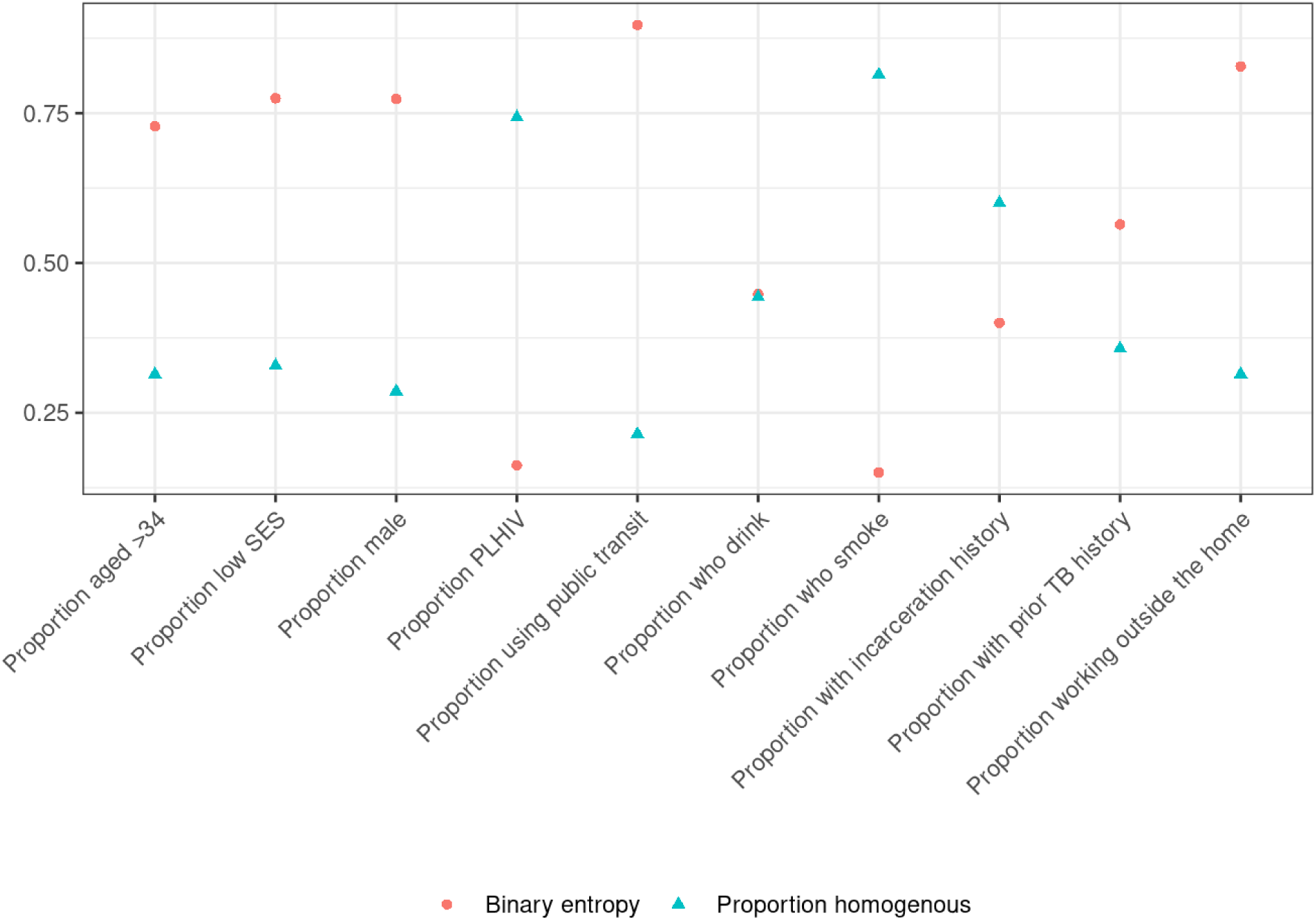
Weighted average binary entropy of clusters and proportion of homogeneous clusters using data from Lima, Peru. Values closer to 0 correspond to higher homogeneity. Features with a large proportion of homogenous clusters would be expected to have lower entropy. Definition of abbreviations: SES = socioeconomic status; PLHIV = people living with HIV.

#### 3.2.1 Comparison to traditional SNP distance cut-off methods

Using an upper bound of 12 and 20 SNPs in the mlTransEpi algorithm results in the same network clusters, whereas SNP distance-based clusters vary by SNP cutoff (Table 1), indicating network-based clusters are more robust to SNP bounds than traditional methods. Our network-based clusters are smaller on average than those created using SNP cutoffs and have a lower average maximum SNP distance (5.0 for Infomap clusters, 6.9 for clusters with SNP cutoff of 12, and 10.8 for clusters with SNP cutoff of 20). Network-based clusters exclude more individuals than SNP based clusters. Individuals are excluded from network-based clusters if they do not have a high probability infector and are excluded from SNP distance-based clusters if they are not within 12 or 20 SNPs of anyone else in the cohort.

**Table 1.** Comparison of clusters using network methods and hierarchical clustering (HC) for edge trimming to traditional clustering methods with varying single nucleotide polymorphism (SNP) distance cutoffs. Unclustered individuals refers to those who were found to not cluster with any other individuals. Note that using an upper bound of 12 and 20 SNPs in the mlTransEpi algorithm results in the same clustered network.

|  | Number of<br>single<br>(unclustered)<br>individuals | Number<br>of<br>clusters | Mean<br>cluster<br>size | Maximum<br>cluster<br>size | Mean<br>maximum<br>SNP<br>distance<br>within<br>cluster |
| --- | --- | --- | --- | --- | --- |
| Network clusters, upper bound = 12 | 1955 | 275 | 3.47 | 45 | 4.95 |
| Network clusters, upper bound = 20 | 1955 | 275 | 3.47 | 45 | 4.95 |
| SNP Clusters, cutoff = 12 | 1072 | 321 | 5.72 | 168 | 6.9 |
| SNP Clusters, cutoff = 20 | 866 | 313 | 6.52 | 174 | 10.78 |

## 4. DISCUSSION

We use network techniques to organize and analyze estimated directed pairwise transmission probabilities generated using mlTransEpi. We compare these clusters to traditional SNP distance-based clusters and show that while our clusters remain invariant under different SNP distance cutoffs in the mlTransEpi algorithm, traditional methods are sensitive to the chosen SNP distance cutoff. We also find that TB transmission clusters in our Lima, Peru settings are heterogenous across most individual level features, potentially indicating significant mixing between age, sex, and SES groups. These methods are useful to better understand transmission dynamics and can be applied to any infectious disease with WGS and/or contact tracing data.

We find high levels of heterogeneity across clusters, particularly by age, sex, SES status, public transit use, and working outside the home. Recent research has begun to deemphasize the role of household TB transmission, particularly in high burden TB settings, and suggest that transmission may not always be between known close contacts.^24, 25^ We would expect this to result in higher rates of assortative mixing amongst those transmitting TB, as seen in our results.

We see high homogeneity in HIV and smoking across clusters, although this may be due to very low prevalences of HIV positivity and smoking in our cohort (Table S2a). Incarceration history also has a low prevalence in our cohort, however it has moderate entropy and proportion of purely homogeneous clusters. Prisons are known TB transmission hotspots,^26, 27^ particularly in Latin America.^28, 29^ Our results indicate that people with a history of incarceration are distributed amongst transmission clusters rather than being clustered together, indicating that they may be contributing to community spread of TB. We also see moderate entropy amongst people who drink; these individuals are contributing to transmission clusters that are both homogenous and heterogenous for drinking status.

Alcohol use is a known risk factor for TB, with multiple outbreaks being traced to people who frequent bars. Our results suggest that these individuals contribute to general community spread, not just amongst those who also drink.

A strength of using mlTransEpi generated probabilities for cluster analysis is that the results do not depend solely on genetic data and/or diagnosis dates. Existing methodologies relying on these data^5, 7, 8^ assume that pathogen genomes mutate rapidly enough to detect differences between infected individuals.^9^ TB has a low mutation rate,^30, 31^ suggesting that methodologies using genetic information alone may not adequately capture transmission dynamics. Furthermore, existing methods often depend on an arbitrary SNP distance cutoff to determine clusters.^9^, ^23^

mlTransEpi is robust to varying SNP cutoffs, and it softens the assumption that SNP distances either do or do not indicate transmission by denoting case pairs as probable (rather than confirmed) transmission events and estimates transmission probabilities for all pairs within its iterative algorithm.^10^ We showed that clusters based on mlTransEpi probabilities are invariant to varying cutoffs, while traditional clusters are not. That said, clusters based on mlTransEpi probabilities also exclude more individuals than traditional SNP based clusters, due to lack of high probability infectors. This is both a strength and a weakness of these network clusters, as only individuals for whom we can identify high probability infectors will be included. Thus, while we cluster fewer individuals, we have a higher confidence in the relationships between individuals in a cluster.

Alternative methods such as TransPhylo^32^ detect clusters on reconstructed phylogenies and transmission trees that are generated via a Bayesian model that incorporates diagnosis dates and genetic sequences. These models are resource intensive and rely on strong evolutionary model assumptions, as the user must provide often unknown parameters to inform the evolutionary model. TransPhylo methods are also extremely sensitive to the proportion of cases sampled.^33^ New work incorporates epidemiological data into the TransPhylo framework, which has improved performance, but the method still remains computationally intensive.^34^ Our methods do not rely on such assumptions, and we demonstrate that our clusters are invariant to multiple sampling scenarios (Supplemental Materials Figure S3). This may be due to mlTransEpi not assuming one true infector for each infectee but rather calculating the relative probabilities for all possible infectors, as well as excluding individuals for which we do not have a high probability infector from clustering analyses. Additionally, we also allow infectors to be diagnosed after infectees. This is important for a disease such as TB, which often has a long and variable diagnostic delay and latent and subclinical period, such that the infectee may develop disease and symptoms before the infector.^35, 36, 37^

Our clusters are based on underlying transmission probabilities generated using naïve Bayes, a simple and well-studied machine learning classifier.^38, 39^ We chose naïve Bayes over more complex machine learning algorithms because of its transparency and ease of use, particularly regarding sparse and missing data.^10^ Though it has many advantages, naïve Bayes assumes that covariates are independent when conditioning on the outcome.

While this assumption may be unrealistic, numerous studies demonstrate that naïve Bayes is robust to violations of this assumption.^38, 39, 40^ Multiple studies have introduced extensions of naïve Bayes for situations in which covariates are dependent.^41, 42^ Future work seeks to incorporate these extensions into mlTransEpi.

While our results were independent of clustering algorithm specifications, the algorithms we used had limitations. The Leiden algorithm is a local community detection algorithm that optimizes modularity to determine clusters.^17^ It builds upon the Louvain method, which was shown to produce poorly connected communities and merge smaller communities into larger ones, by employing a refinement phase that allows communities to be split in order to ensure all communities are well-connected.^17^ That said, it can miss small clusters in certain cases, and relies on specification of a resolution parameter for which there is no one optimal value.^17^ While the Infomap algorithm does not require specification of a resolution parameter, it too has biases. It sometimes combines multiple clusters into a single community if edges between them have enough weight to guide the random walk or if nodes exist with high edge weights to multiple other clusters.^16, 43^ These two algorithms were selected because they are well researched and perform on weighted, directed networks. Future work seeks to examine and apply other network clustering algorithms for transmission cluster detection.

We demonstrate a method to organize pairwise directed transmission probabilities into transmission clusters and analyze these clusters to determine which demographics tend to cluster together. Both the underlying transmission probabilities and clustering algorithms rely on minimal assumptions, making this a data-driven method that can be applied to any infectious disease outbreak dataset with demographic data, WGS, and/or contact tracing data. An understanding of disease transmission clusters can be used to inform disease and targeted interventions to interrupt transmission.

## DECLARATIONS

### Ethics approval

The Harvard School of Public Health institutional review board and Peru’s Research Ethics Committee of the National Institutes of Health gave ethical approval for this work. All study participants provided voluntary written informed consent prior to study participation.

## Data Availability

Data cannot be shared publicly to protect study participant privacy.

## Acknowledgements

The authors thank Leonid Lecca, Roger I. Calderon, Carmen C. Contreras, Judith Jimenez, and others at Socios en Salud in Lima, Peru. We also thank the patients, their families, and the healthcare personnel at the participating health centers in Lima, Peru. Without these people, this study would not have been possible.

## Author contributions

ANS, HEJ, and LFW designed the study methodology. ANS implemented the analysis and drafted the manuscript. MBB, MBM and CCH advised study implementation. All authors aided in manuscript revision.

## Supplemental materials

Supplementary materials are available at *IJE* online

## Data availability

Data cannot be shared publicly to protect study participant privacy. Code is available upon request.

## Use of Artificial intelligence (AI) tools

No AI tools were used in this analysis or drafting of this manuscript.

