## Supplemental Materials for "Network Analysis of Pairwise Relative Tuberculosis Transmission Probabilities in Lima, Peru"

**Table of Contents**

**S1. Simulation studies**......................................................................................................2

**S2. Peru data results**.........................................................................................................14

**Section S1. Edge trimming methods**

Probability cutoff: We remove all edges with a weight below a pre-specified probability.

Hierarchical clustering: We consider infectors to be high probability infectors if the gap between the two estimated average, unscaled transmission probabilities is greater than some cutoff.

Kernel density estimation: we use a simple rectangular kernel and consider infectors to be high probability infectors if the kernel density estimate equals 0 at some value within the range of estimated transmission probabilities and their estimated average probability is greater than the lowest probability for which the binwidth equals 0.

We assess edge removal method performance with varying binwidths and cutoffs via simulation study (described in detail in Main Text section 2.3).

### Section S2. Simulation studies

#### Outbreak simulation

We simulate outbreaks consisting of multiple transmission chains and their corresponding phylogenetic trees via R package TransPhylo v1.4.5[18] and generate genetic sequences for the phylogenetic trees using the R package phangorn v 2.11.1.[19] The simulations assume a reproductive number of 1.2, a gamma-distributed generation interval with a shape parameter of 1.2, scale parameter of 2, and shift parameter of 0.25[20] (mean generation interval of 2.65 years), and a mutation rate of 0.5 SNPs per genome per year.[21] We vary the generation interval distribution in sensitivity analyses. We allow simulations to run for 20 years. TransPhylo simulates observation dates to better reflect the timing between onset of disease and observation by building in a sampling distribution. We assume the same distribution for the generation interval and sampling distribution.

#### Covariate construction details

We simulate four covariates, $X_i,\ i = 1, \dots, 4$, at the individual level based on the outbreak's phylogenetic tree. Each of the simulated transmission chains has a source case for whom we first assign a value according to the values in the second column of Table S1. We then assign each subsequent individual's variable value according to the values specified in the third column of Table S1. Finally, we construct pairwise variables as specified in the fourth column of Table S1.

**Table S1**. Simulated covariates structures. $X_i$ refers to the individual level covariate and $Z_i$ refers to the pair level covariate. Source variable frequency refers to the frequency for the starts of transmission chains. Linked variable frequency refers to the frequency for all other individuals.

| Variable | Source individual level variable frequency | Linked individual level variable frequency | Paired variable construction | Motivating Example |
|---|---|---|---|---|
| $X_1/Z_1$ | 0: 60%; 1: 40% | *If* $X_1 = X_5 = 0$*:*<br>80% chance of same<br>20% chance of different<br>*If* $X_1 = X_5 = 1$*:*<br>20% chance of same<br>80% chance of different<br>*If* $X_1 \neq X_5$*:*<br>50% chance of same<br>50% chance of different | $Z_1 = 1$ if same<br>$Z_1 = 0$ if different | Sex |
| $X_2/Z_2$ | a: 50%; b: 30%;<br>c: 15%; d:5% | 70% chance of same<br>30% chance of different | $Z_2 = 1$ if same<br>$Z_2 = 0$ if different | Nationality |
| $X_3/Z_3$ | 0: 70%; 1: 30% | *Infector is a:*<br>80% of a-a; 20% of a-b<br>*Infector is b:*<br>70% of b-b; 30% of b-a | $Z_3 = 1$ if 0-0<br>$Z_3 = 2$ if 1-1<br>$Z_3 = 3$ if 0-1<br>$Z_3 = 4$ if 1-0 | Age group |
| $X_4/Z_4$ | a: 5%; b: 5%;<br>c: 5%; d:20%;<br>e: 30%; f:10%;<br>g:10%; h: 5%;<br>i:5%; j:5% | 60% chance of same<br>35% chance of neighbors<br>5% chance of other | $Z_4 = 1$ if same<br>$Z_4 = 2$ if neighbors<br>$Z_4 = 3$ otherwise | Country of origin |

**Clustering metric details**

Mutual information (MI) is a measure of the dependence of one random variable (the true clusters) on another (the estimated clusters). It has a lower bound of 0 and no upper bound.

The pairwise F score and Fowlkes-Mallows (FM) index are calculated by considering the estimated and true cluster assignments of all pairwise combinations of nodes. The pairwise F-score is calculated as

$$F = 2\frac{precision * recall}{precision + recall} = \frac{2TP}{2TP + FP + FN},$$

where TP are true positives (a pair is in both the same estimated and true clusters), FP are false positives (a pair is in the same estimated cluster but not the same true cluster), and FN are false negatives (a pair is in the same true cluster but not the same estimated cluster). It is bounded between 0 and 1, with a score of 1 indicating perfect precision and recall and a score of 0 indicating either precision or recall is 0. The Fowlkes-Mallows (FM) index is calculated as

$$FM = \sqrt{PPV * TPR} = \sqrt{\frac{TP}{TP + FP} * \frac{TP}{TP + FN}},$$

where TPR refers to the true positive rate and TP, FP, and FN are described above. It is also bounded between 0 and 1, with a score of 0 being the worst possible binary classification (i.e. all elements are misclassified) and 1 is the best possible classification.

**Figure S1a**. Edge trimming metrics from 100 simulated outbreaks, using the entire (20 years) outbreak. We consider four edge trimming methods: hierarchical clustering (HC), kernel density (KD) estimation, and probability cutoffs.

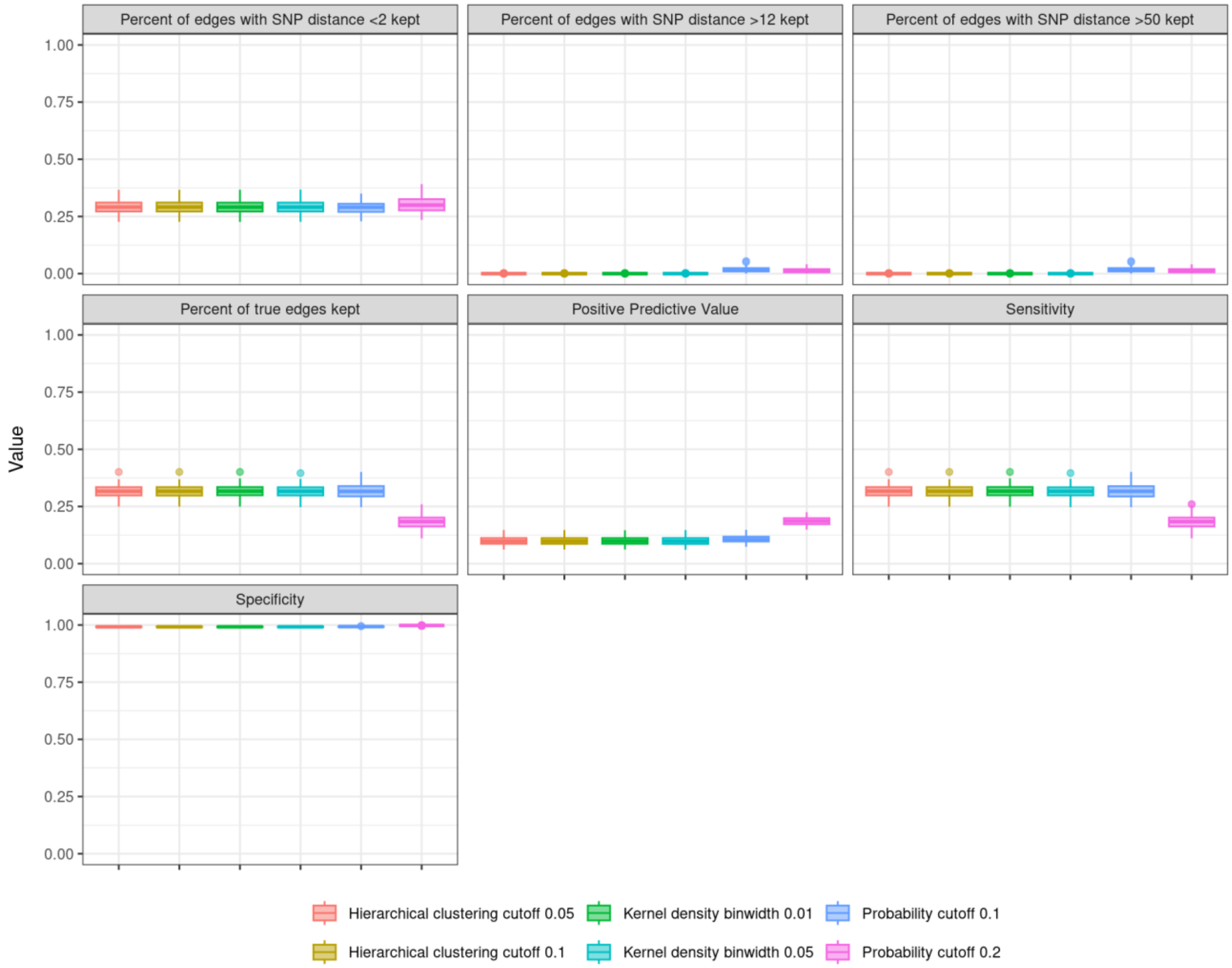

**Figure S1b.** Pairwise F (F), Fowlkes-Mallows (FM), and mutual information (MI) scores assessing clustering performance for 100 simulated outbreaks, using the entire (20 years) outbreak.

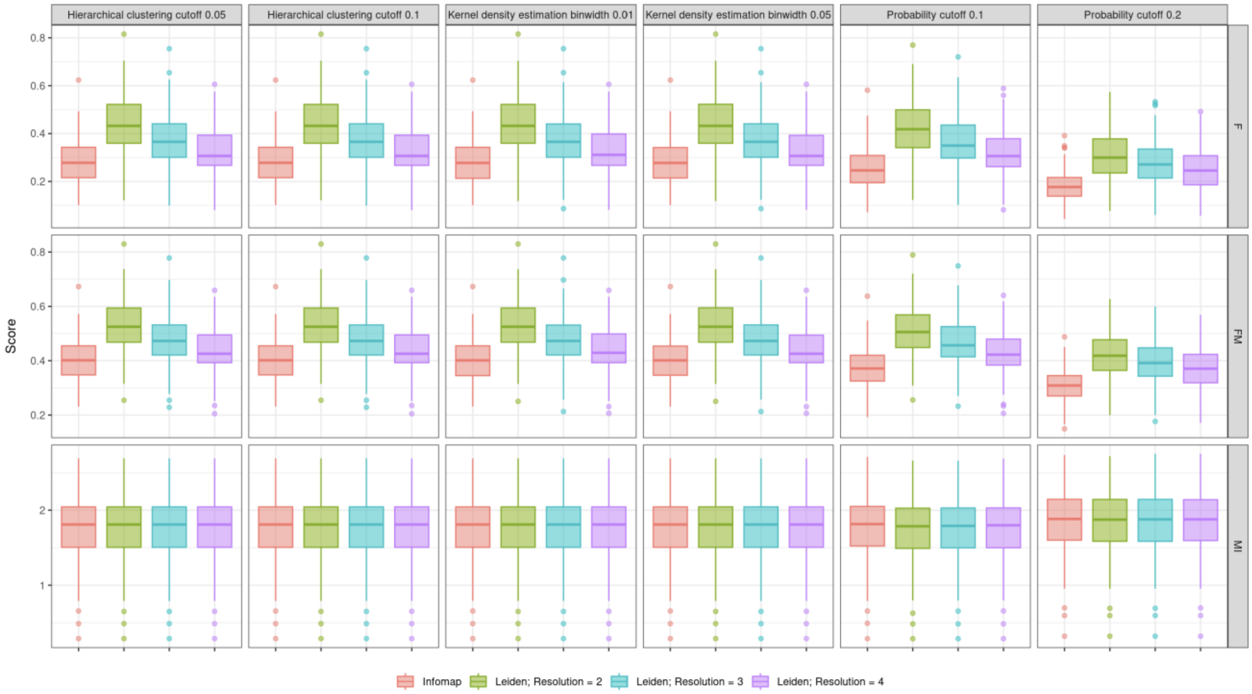

**Figure S2a**. Edge trimming metrics from 100 simulated outbreaks, using the trimmed (8 years) outbreak stratified by stratified by single nucleotide polymorphisms (SNP) cutoff boundaries for the underlying mlTransEpi algorithm. We consider three edge trimming methods: hierarchical clustering (HC), kernel density (KD) estimation, and probability cutoffs. Panel (a) uses a lower bound cutoff of 2 and an upper bound cutoff of 12, panel (b) uses a lower bound cutoff of 4 and an upper bound cutoff of 12, and panel (c) uses a lower bound cutoff of 2 and an upper bound cutoff of 20.

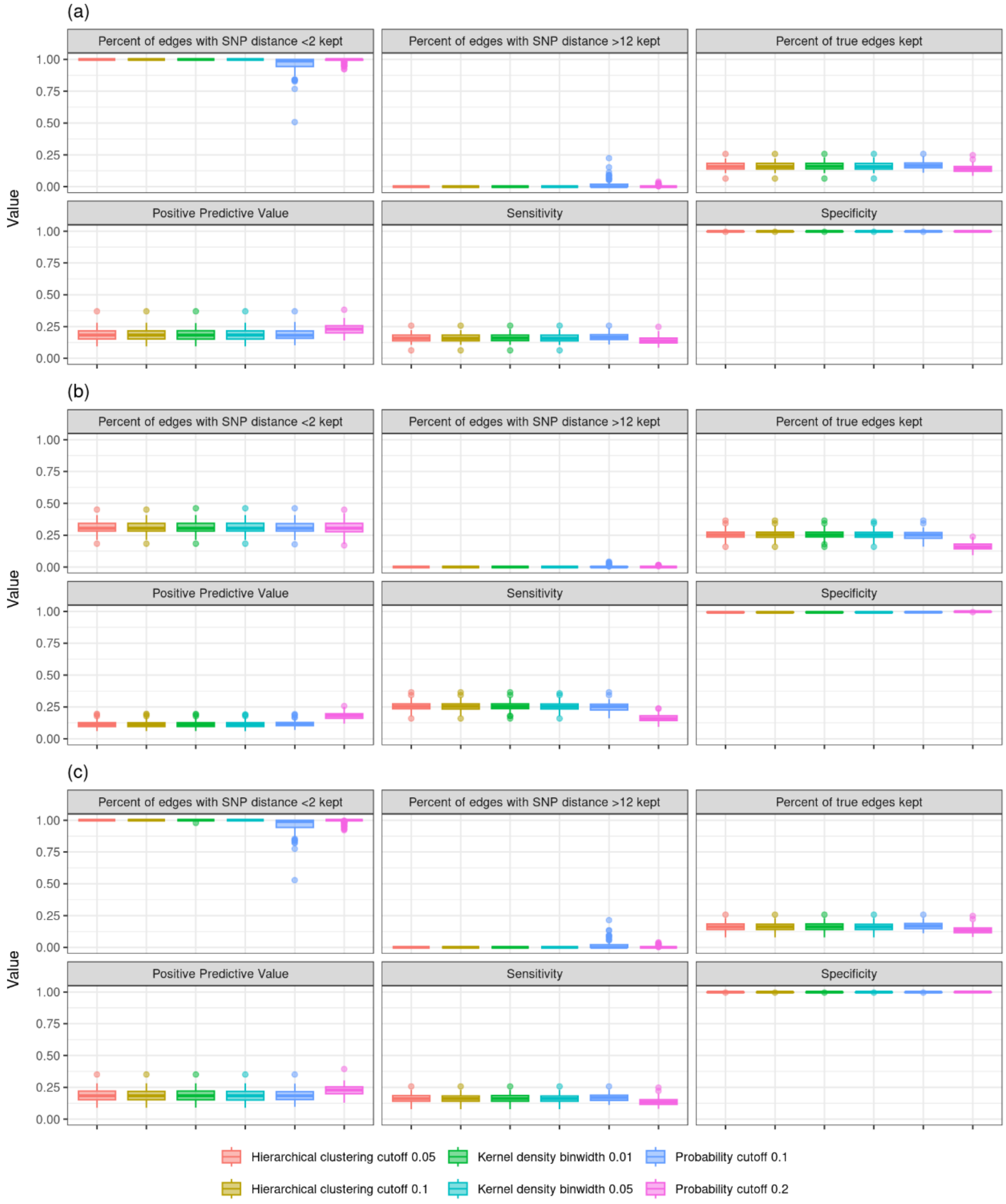

**Figure S2b.** Pairwise F (F), Fowlkes-Mallows (FM), and mutual information (MI) scores assessing clustering performance for 100 simulated outbreaks, using the using the trimmed (8 years) outbreak stratified by stratified by single nucleotide polymorphisms (SNP) cutoff boundaries for the underlying mlTransEpi algorithm. As results are shown to be consistent across edge trimming methods in Figure S6, we only show for hierarchical clustering with a cutoff of 0.1.

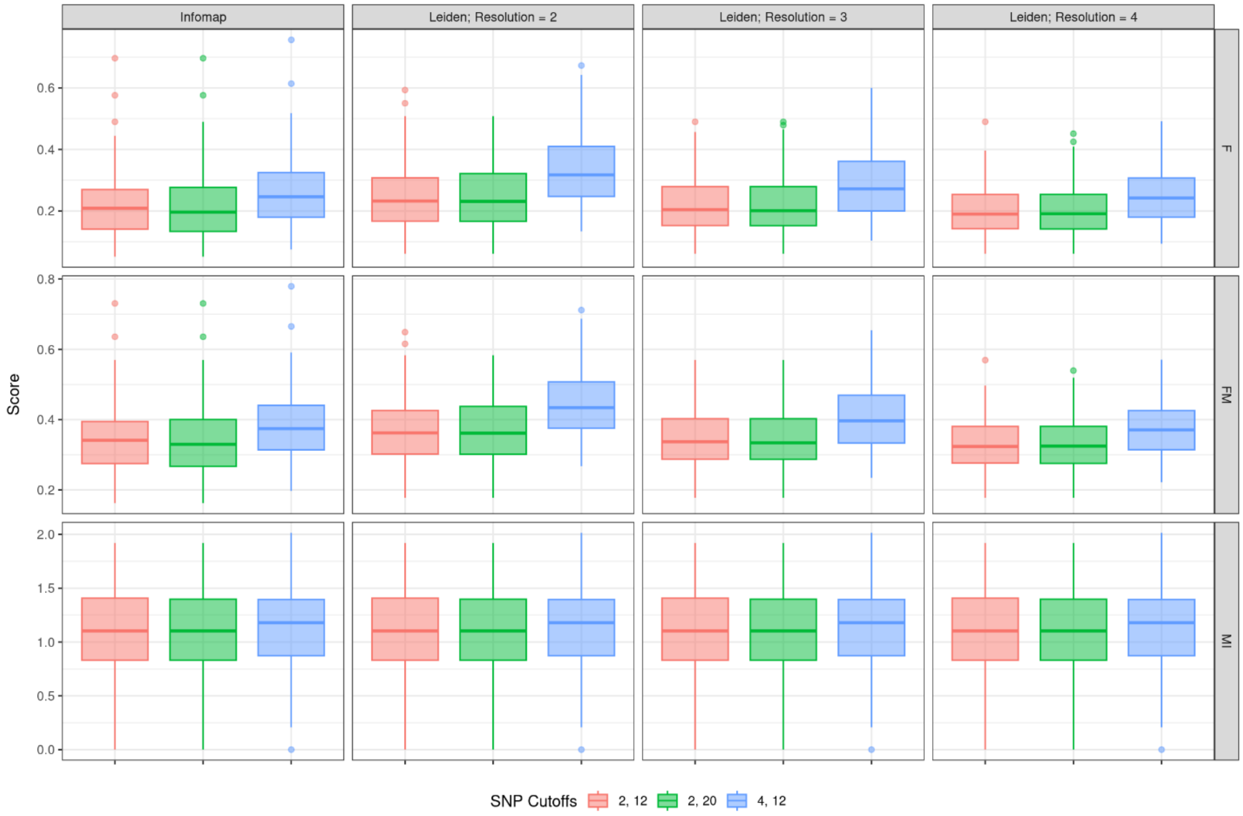

**Figure S3a**. Edge trimming metrics from 100 simulated outbreaks, using the trimmed (8 years) outbreak stratified by stratified by scale parameter used in simulation generation interval. We consider three edge trimming methods: hierarchical clustering (HC), kernel density (KD) estimation, and probability cutoffs. Panel (a) uses a scale parameter of 1.5, panel (b) uses a scale parameter of 2, and panel (c) uses a scale parameter of 2.5.

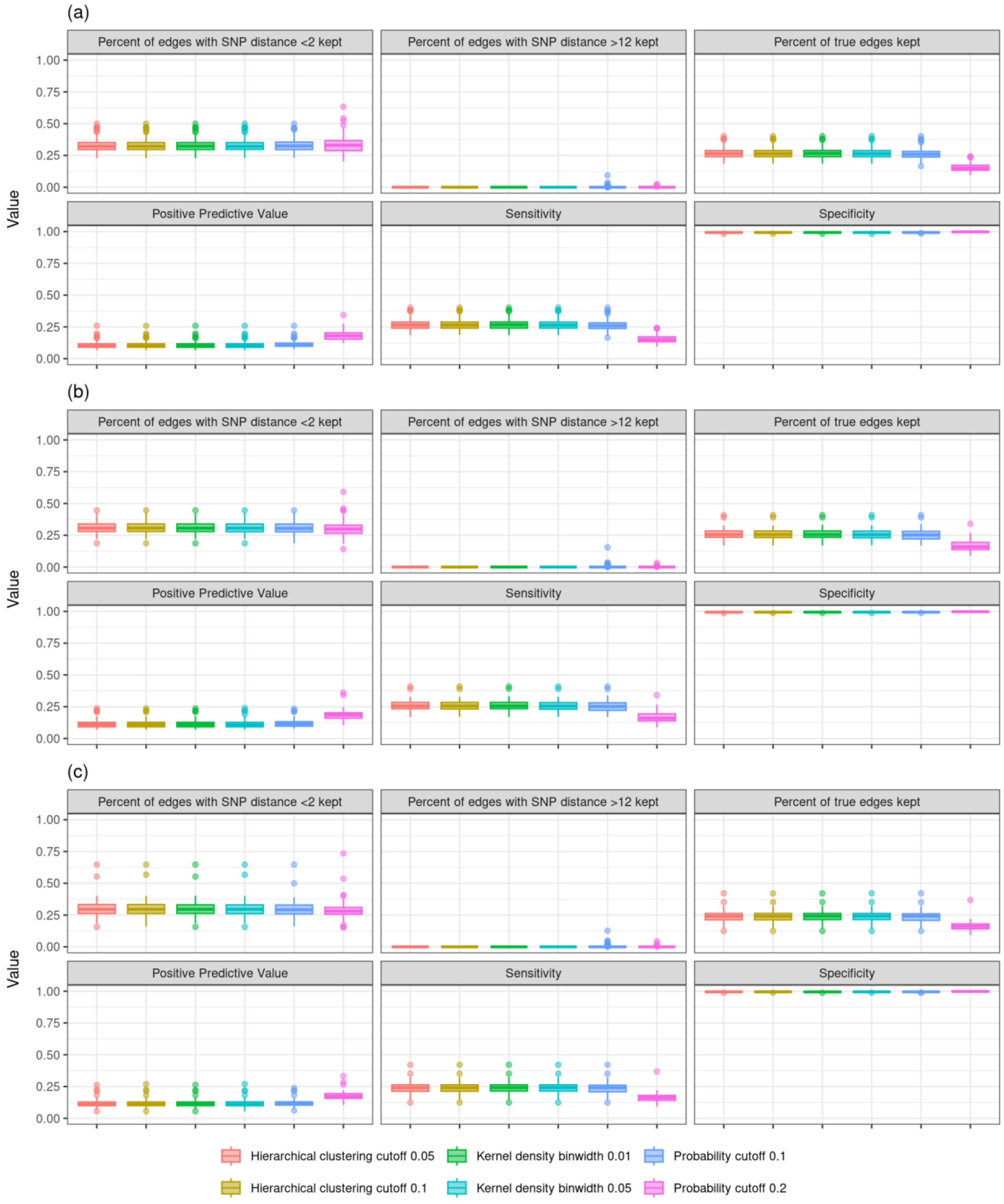

**Figure S3b.** Pairwise F (F), Fowlkes-Mallows (FM), and mutual information (MI) scores assessing clustering performance for 100 simulated outbreaks, using the using the trimmed (8 years) outbreak stratified by stratified by scale parameter used in simulation generation interval (GI). As results are shown to be consistent across edge trimming methods in Figure S6, we only show for hierarchical clustering with a cutoff of 0.1.

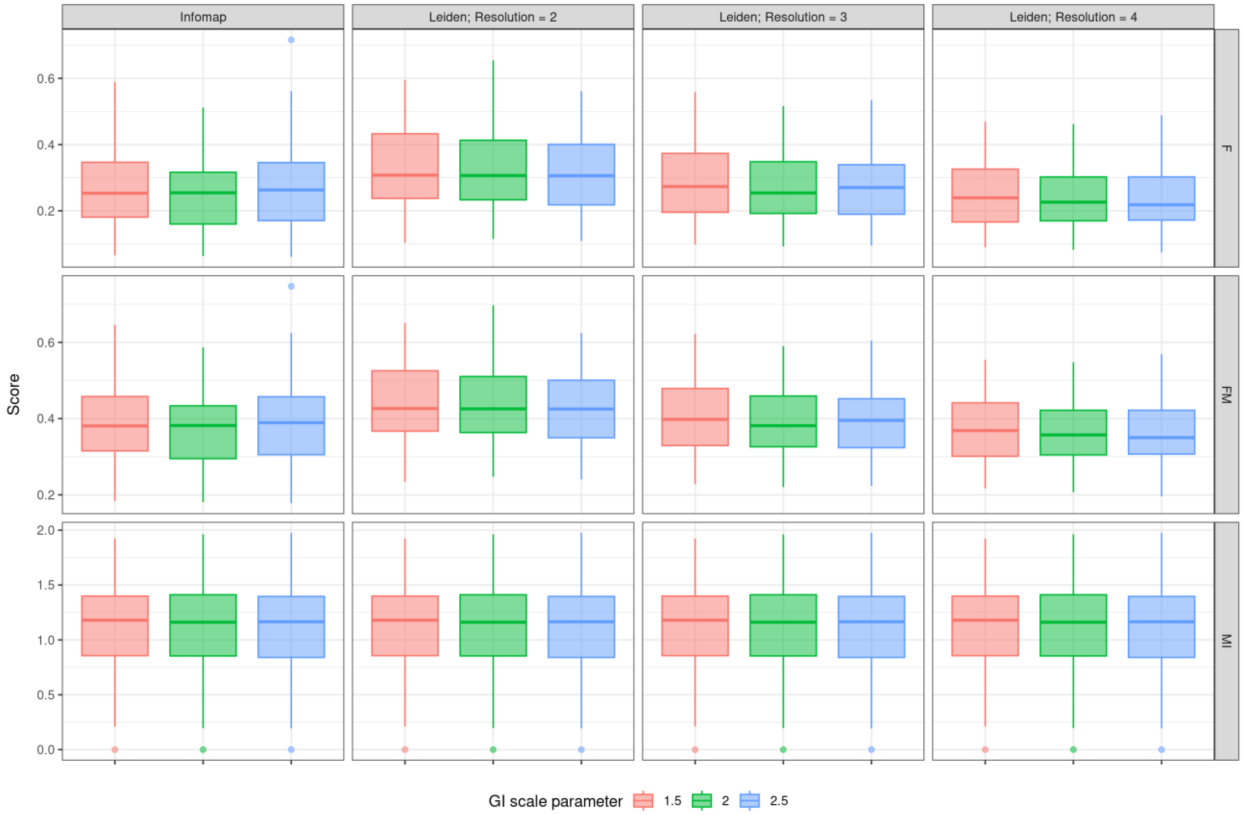

**Figure S4.** Pairwise F (F), Fowlkes-Mallows (FM), and mutual information (MI) scores assessing clustering performance for 100 simulated outbreaks, using the using the trimmed (8 years) outbreak. We vary the amount of time we allow the infector to be diagnosed after the infectee (overlap time). Edges are trimmed using hierarchical clustering with a cutoff of 0.1.

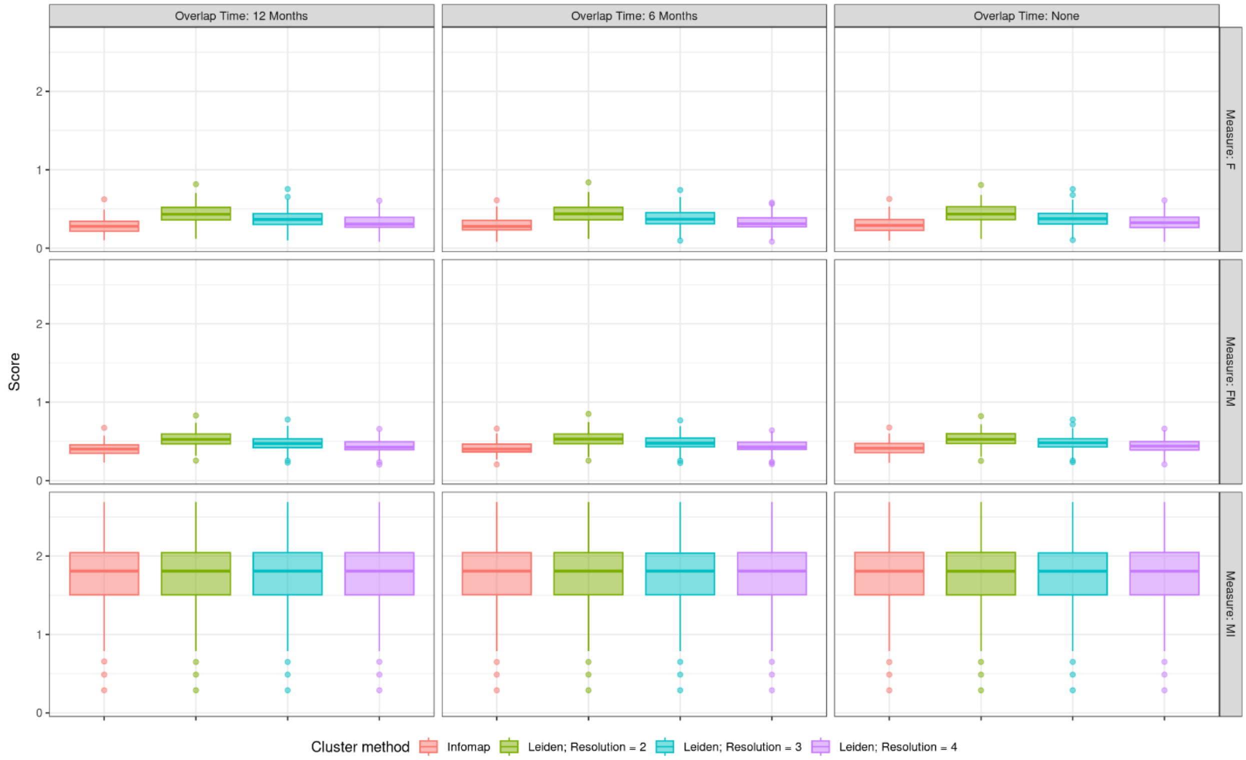

**Figure S5**. Pairwise F (F), Fowlkes-Mallows (FM), and mutual information (MI) scores assessing clustering performance for 100 simulated outbreaks from which cases were sampled according to various schemes, using the trimmed (8 years) outbreak. Edges are trimmed using hierarchical clustering with a cutoff of 0.1.

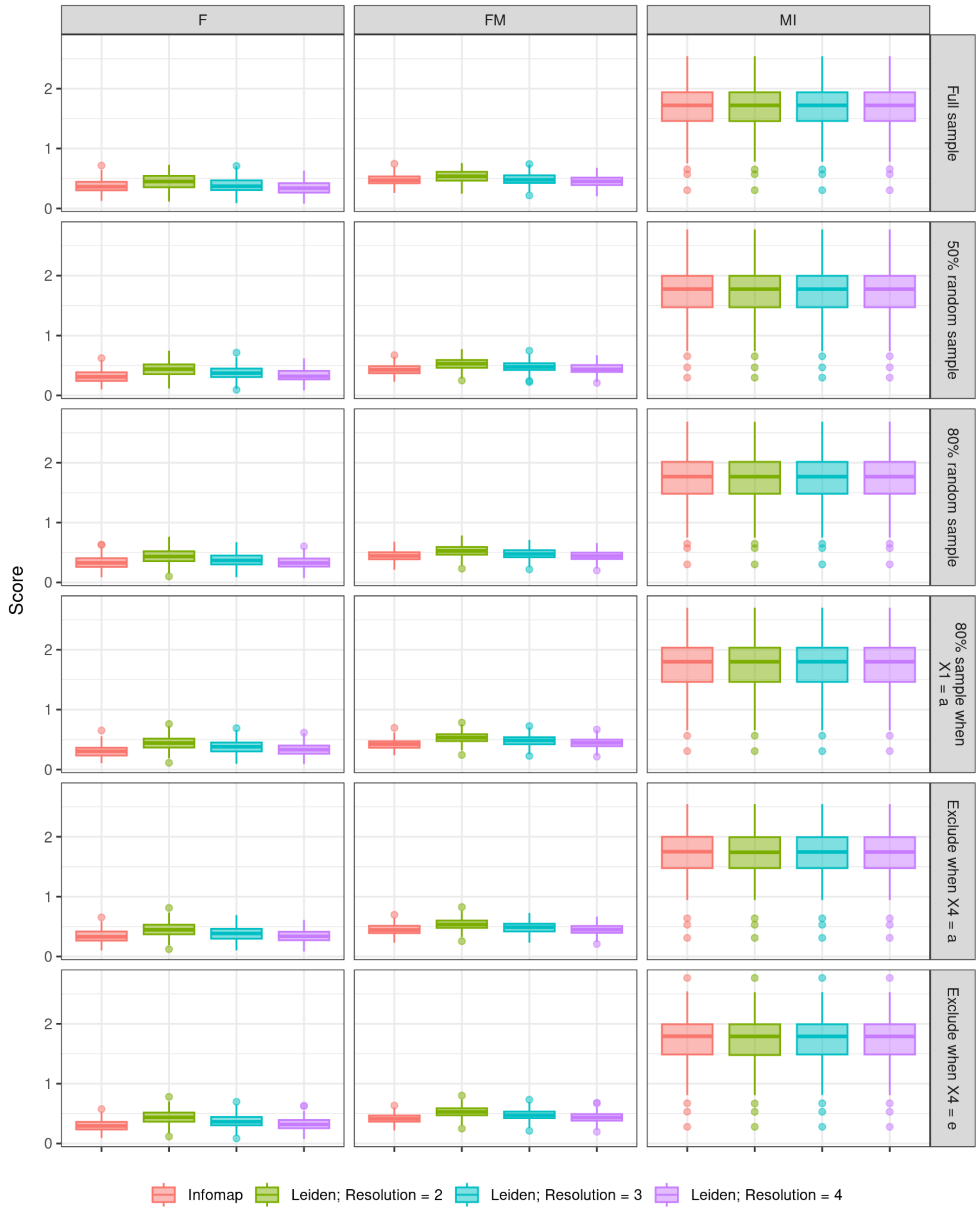

### Section S3. Peru data tabulations

**Table S2a**. Individual level demographic and clinical characteristics

| Variable | Value | Count |
|---|---|---|
| Age | <=34 | 1614 |
| | >34 | 874 |
| Sex | F | 968 |
| | M | 1520 |
| Work outside the home | No | 1552 |
| | Yes | 936 |
| Public transit use | Heavy | 1140 |
| | Light | 742 |
| | Non-public transit user | 606 |
| Socioeconomic status | Low | 822 |
| | Middle | 845 |
| | High | 821 |
| Spent at least one night in prison | No | 2372 |
| | Yes | 116 |
| Smoking status | Heavy | 44 |
| | Light | 20 |
| | Non-Smoker | 2424 |
| Drinking status | Heavy | 268 |
| | Light | 812 |
| | Non-Drinker | 1408 |
| HIV status | Negative | 2401 |
| | Positive | 87 |
| Cough more than 30 days | No | 1231 |
| | Yes | 1257 |
| Prior TB history | No | 2057 |
| | Yes | 431 |
| Smear status | 0 | 622 |
| | 1 | 712 |
| | 2+ | 1154 |

**Table S2b**. Cohort pair-level demographic and clinical characteristics, n (%) stratified by single nucleotide polymorphisms (SNP) differences

| Variable | Level | Total pairs (n =3,078,413) | Pairs with SNP Differences < 4 (n = 2,481) | Pairs with SNP Differences > 12 (n = 3,069,784) |
|---|---|---|---|---|
| Age (years) | >34 → >34 | 382,197 (12.33) | 182 (7.28) | 381,366 (12.34) |
| | >34 → ≤ 34 | 724,002 (23.36) | 450 (17.99) | 722,146 (23.36) |
| | ≤ 34 → >34 | 689,282 (22.24) | 514 (20.55) | 687,516 (22.24) |
| | ≤ 34 → ≤ 34 | 1,304,188 (42.08) | 1,355 (54.18) | 1,299,941 (42.06) |
| Sex | F → F | 468,913 (15.13) | 274 (10.96) | 468,002 (15.14) |
| | F → M | 739,832 (23.87) | 529 (21.15) | 738,004 (23.88) |
| | M → F | 734,330 (23.69) | 580 (23.19) | 732,347 (23.69) |
| | M → M | 1,156,594 (37.31) | 1,118 (44.7) | 1,152,616 (37.29) |
| Work outside the home | No → No | 2,123,324 (68.5) | 1,504 (60.14) | 2,117,852 (68.52) |
| | No → Yes | 295,313 (9.53) | 302 (12.08) | 294,410 (9.52) |
| | Yes → No | 218,427 (7.05) | 231 (9.24) | 217,709 (7.04) |
| | Yes → Yes | 265,210 (8.56) | 265 (10.6) | 264,298 (8.55) |
| Public transit use | Heavy → Light | 197,395 (6.37) | 199 (7.96) | 196,700 (6.36) |
| | Light → Heavy | 1,205,891 (38.9) | 1,022 (40.86) | 1,202,553 (38.91) |
| | Light → Light | 714,371 (23.05) | 591 (23.63) | 712,412 (23.05) |
| Socioeconomic status | Low → Low | 740,958 (23.9) | 578 (23.11) | 738,797 (23.9) |
| | Low → Not low | 438,449 (14.15) | 310 (12.4) | 437,207 (14.14) |
| | Not low → Low | 650,464 (20.98) | 531 (21.23) | 648,565 (20.98) |
| | Not low → Not low | 755,616 (24.38) | 648 (25.91) | 753,451 (24.38) |
| Spent at least one night in prison | No → No | 783,987 (25.29) | 619 (24.75) | 781,795 (25.29) |
| | No → Yes | 909,602 (29.35) | 703 (28.11) | 907,158 (29.35) |
| | Yes → No | 338,059 (10.91) | 325 (12.99) | 336,911 (10.9) |
| | Yes → Yes | 727,250 (23.46) | 547 (21.87) | 725,170 (23.46) |
| Smoking status | Nonsmoker → Nonsmoker | 644,737 (20.8) | 512 (20.47) | 642,837 (20.8) |
| | Nonsmoker → Smoker | 1,389,623 (44.83) | 1,117 (44.66) | 1,386,051 (44.84) |
| | Smoker → Nonsmoker | 2,817,319 (90.89) | 1,824 (72.93) | 2,810,372 (90.92) |
| | Smoker → Smoker | 136,613 (4.41) | 256 (10.24) | 135,864 (4.4) |
| Drinking status | Nondrinker → Nondrinker | 139,056 (4.49) | 343 (13.71) | 138,198 (4.47) |

| | | | | |
|---|---|---|---|---|
| | Nondrinker → Drinker | 6,681 (0.22) | 78 (3.12) | 6,535 (0.21) |
| | Drinker → Nondrinker | 2,942,228 (94.92) | 2,313 (92.48) | 2,934,159 (94.93) |
| | Drinker → Drinker | 75,525 (2.44) | 64 (2.56) | 75,268 (2.44) |
| HIV status | Negative → Negative | 79,896 (2.58) | 115 (4.6) | 79,540 (2.57) |
| | Negative → Positive | 2,020 (0.07) | 9 (0.36) | 2,002 (0.06) |
| | Positive → Negative | 992,448 (32.02) | 582 (23.27) | 990,348 (32.04) |
| | Positive → Positive | 704,617 (22.73) | 524 (20.95) | 702,621 (22.73) |
| Diabetes | No → No | 818,886 (26.42) | 729 (29.15) | 816,529 (26.42) |
| | No → Yes | 583,718 (18.83) | 666 (26.63) | 581,471 (18.81) |
| | Yes → Everyone else | 2,886,663 (93.13) | 2,340 (93.56) | 2,878,528 (93.13) |
| Cavity | No → Everyone else | 99,000 (3.19) | 69 (2.76) | 98,740 (3.19) |
| | Yes → Everyone else | 110,259 (3.56) | 86 (3.44) | 109,967 (3.56) |
| Cough | No → Everyone else | 3,747 (0.12) | 6 (0.24) | 3,734 (0.12) |
| | Yes → Everyone else | 2,762,968 (89.14) | 2,310 (92.36) | 2,754,969 (89.13) |
| Prior TB history | No → Everyone else | 163,395 (5.27) | 90 (3.6) | 163,011 (5.27) |
| | Yes → Everyone else | 173,306 (5.59) | 101 (4.04) | 172,989 (5.6) |
| Smear | Negative → Everyone else | 2,226,508 (71.83) | 1,755 (70.17) | 2,220,287 (71.83) |
| | Positive → Everyone else | 873,161 (28.17) | 746 (29.83) | 870,682 (28.17) |

**Table S3.** Network characteristics by varying hierarchical clustering cutoff. All possible edges and all possible nodes refer to the number of edges and nodes before hierarchical clustering was applied. Note that cutoffs of 0.05 and 0.1 result in the same network. Network and cluster characteristics are similar across cutoffs.

| Value | Cutoff = 0.05 | Cutoff = 0.1 | Cutoff = 0.15 | Cutoff = 0.2 |
|---|---|---|---|---|
| N edges (% all possible edges) | 2380 (0.1%) | 2380 (0.1%) | 1682 (0.1%) | 1647 (0.1%) |
| N nodes (% all possible nodes) | 954 (32.8%) | 954 (32.8%) | 932 (32%) | 930 (32%) |
| N disconnected subgraphs | 268 | 268 | 268 | 268 |
| N Infomap clusters | 275 | 275 | 278 | 278 |
| N Leiden resolution = 1 clusters | 267 | 267 | 267 | 267 |
| N Leiden resolution = 3 clusters | 268 | 268 | 268 | 268 |
| N Leiden resolution = 5 clusters | 269 | 269 | 268 | 268 |
| N subgraphs of one Infomap cluster (% of subgraphs) | 262 (97.8%) | 262 (97.8%) | 262 (97.8%) | 262 (97.8%) |
| N subgraphs of one Leiden resolution = 1 cluster (% of subgraphs) | 268 (100%) | 268 (100%) | 268 (100%) | 268 (100%) |
| N subgraphs of one Leiden resolution = 3 cluster (% of subgraphs) | 267 (99.6%) | 267 (99.6%) | 267 (99.6%) | 267 (99.6%) |
| N subgraphs of one Leiden resolution = 5 cluster (%of subgraphs) | 266 (99.3%) | 266 (99.3%) | 267 (99.6%) | 267 (99.6%) |

**Figure S6**. Binary entropy of clusters using data from Lima, Peru. Clusters are generated using a hierarchical clustering cutoff 0f 0.1 and Infomap clustering. Values closer to 0 correspond to higher homogeneity.

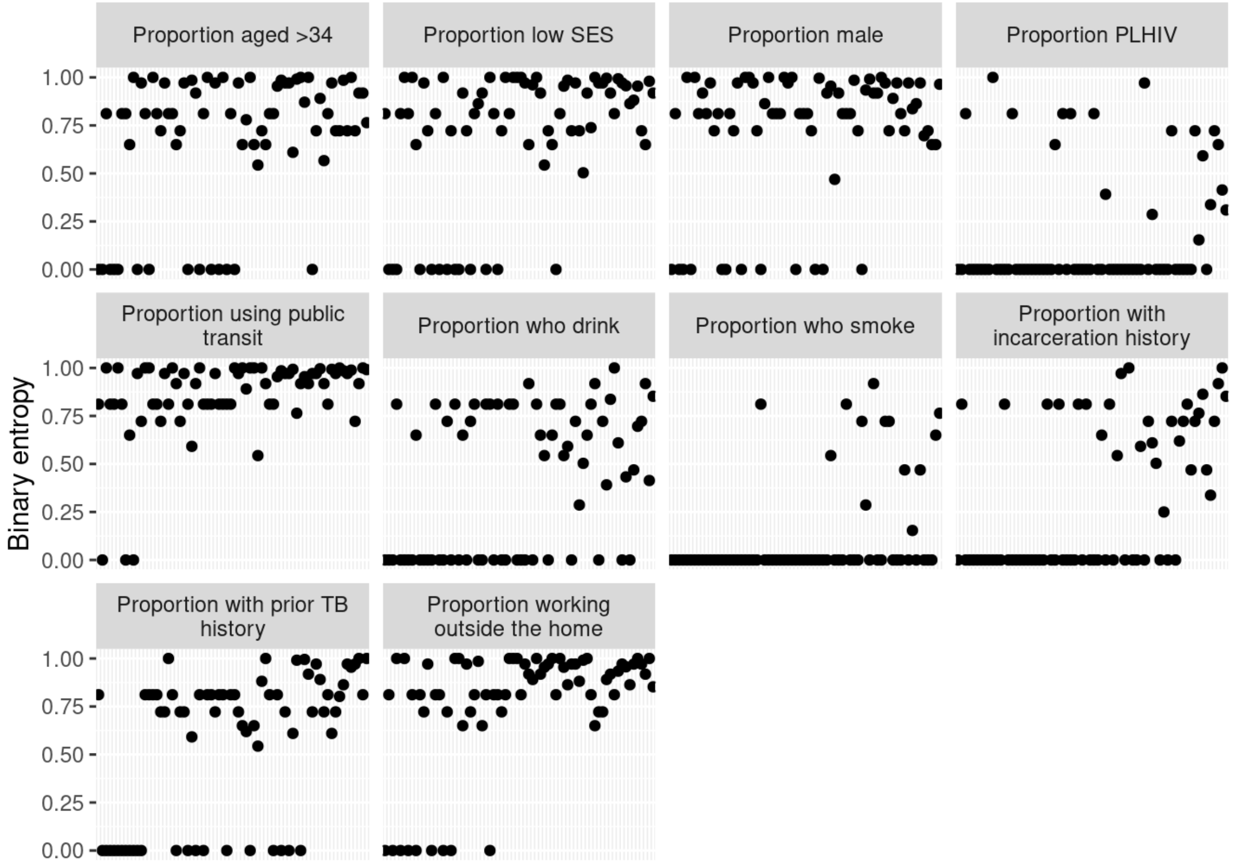

**Figure S7.** Weighted average binary entropy of clusters using data from Lima, Peru. (a) presents results from a graph with trimmed edges using hierarchical clustering with a cutoff of 0.05, (b) with a cutoff of 0.1, (c) with a cutoff of 0.15, and (d) with a cutoff of 0.2. Values closer to 0 correspond to higher homogeneity. Results are consistent across clustering methods.

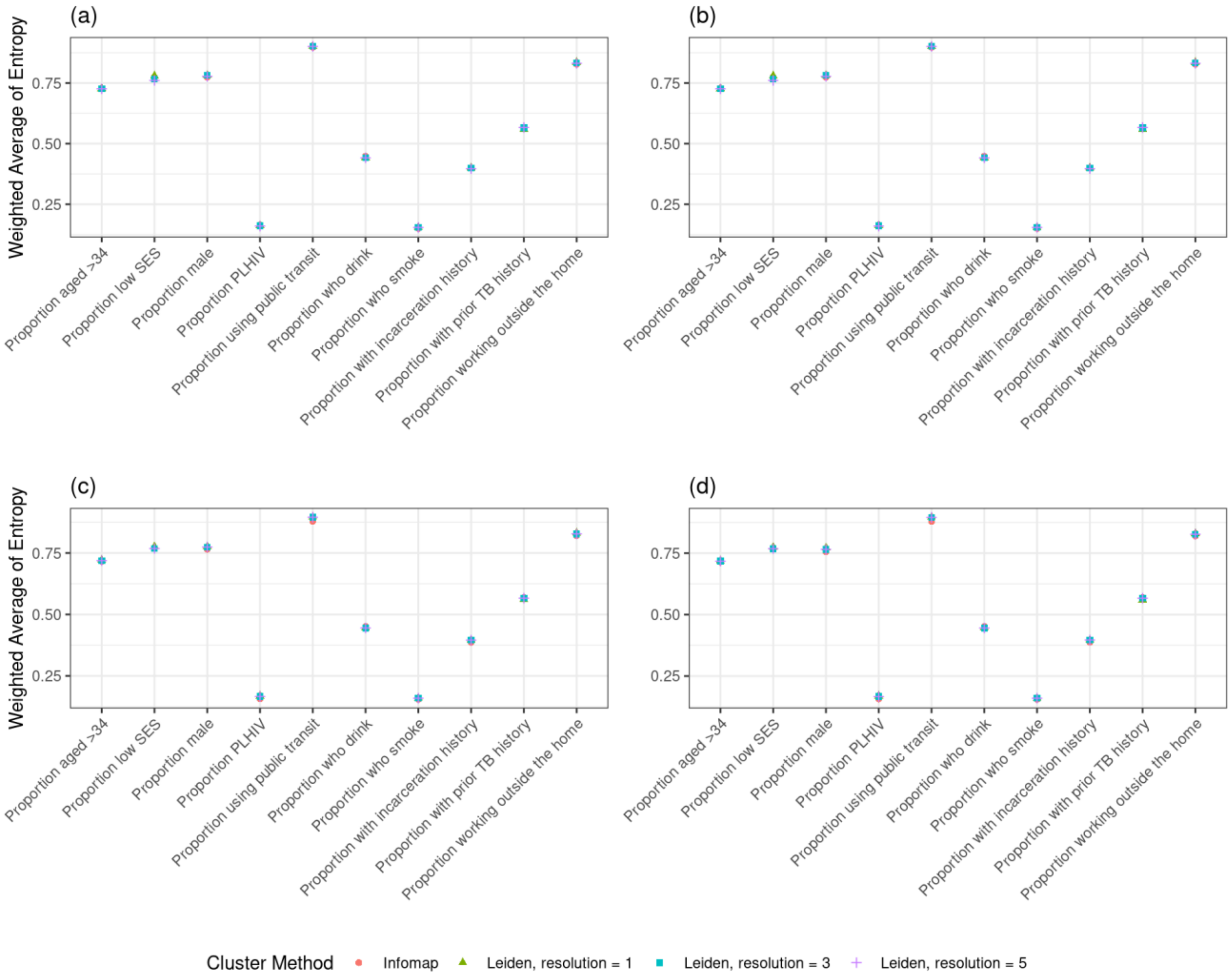

**Figure S8.** Proportion of fully homogeneous clusters using data from Lima, Peru. (a) presents results from a graph with trimmed edges using hierarchical clustering with a cutoff of 0.05, (b) with a cutoff of 0.1, (c) with a cutoff of 0.15, and (d) with a cutoff of 0.2. Results are consistent across clustering methods.

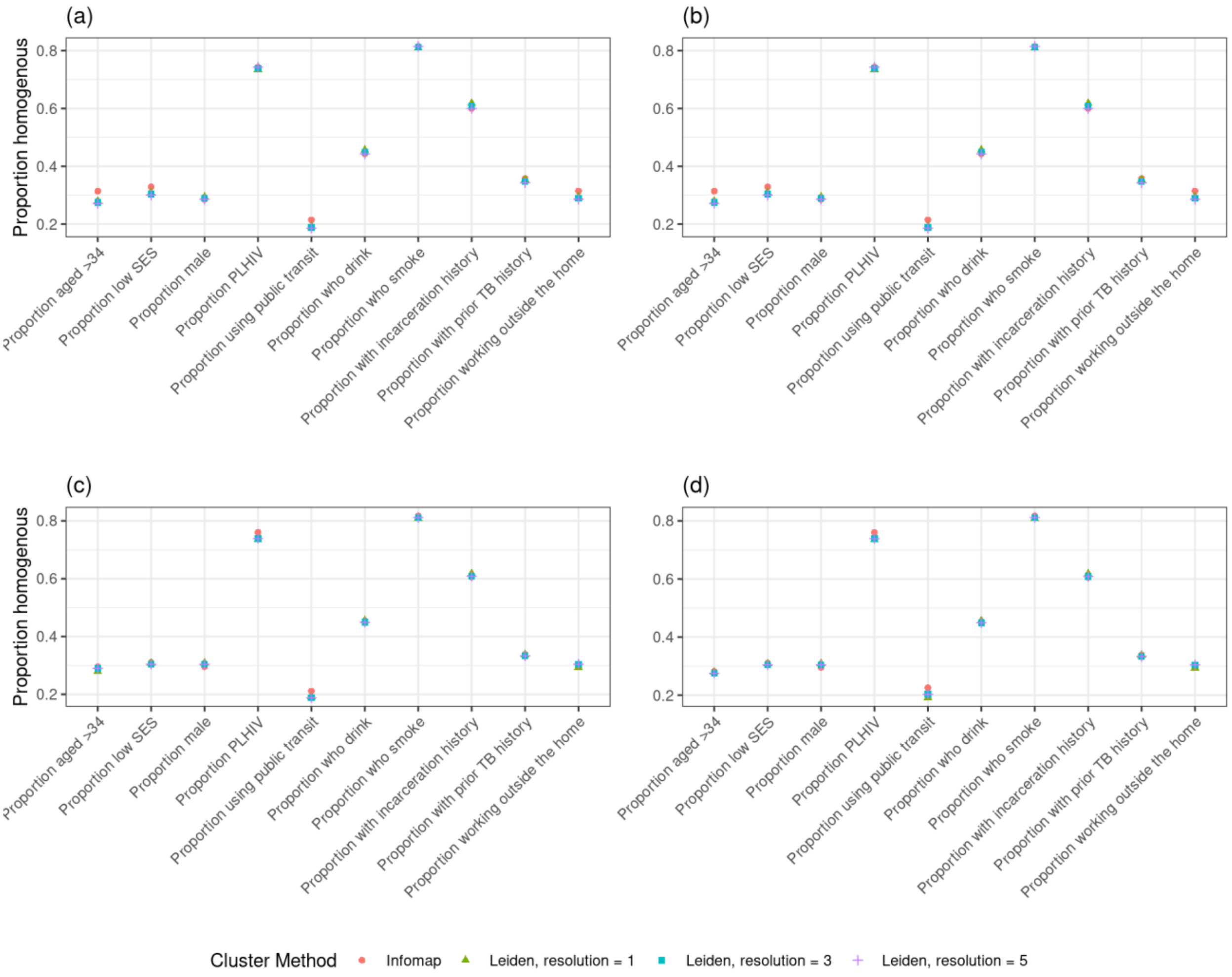
